# Cumulative risk of revision after primary total hip arthroplasty in registries: Systematic review and meta-analysis of selected hip stems and cups

**DOI:** 10.1101/2024.04.03.24305257

**Authors:** Christophe Combescure, James A Smith, Christophe Barea, Lotje A. Hoogervorst, Rob Nelissen, Perla J Marang-van de Mheen, Anne Lübbeke, the arthroplasty registry group

**Affiliations:** Division of Clinical Epidemiology, Geneva University Hospitals, Switzerland; Nuffield Department of Orthopaedics, Rheumatology and Musculoskeletal Sciences, University of Oxford, United Kingdom; Division of Orthopaedic Surgery, Geneva University Hospitals, Switzerland; Department of Orthopaedics, Leiden University Medical Center, Netherlands; Safety & Security Science and Centre for Safety in Healthcare, Delft University of Technology, Delft, Netherlands

**Keywords:** total hip arthroplasty, registry, revision, implant, systematic review, meta-analysis

## Abstract

**Purpose:** The objective was to investigate the consistency in cumulative revision rates for a selection of total hip arthroplasty cups and stems across national/regional hip arthroplasty registries worldwide.

**Methods:** Ten cups and 10 stems from total hip systems were randomly selected. Two frequently used implants across registries were added, totalling 11 cups and 11 stems. Cumulative revision rates (CRR) and 95%CIs were extracted from the latest annual registry reports using these implants. CRRs were pooled for each cup or stem, and differences between cup-stem combinations and between registries were investigated.

**Results:** CRRs were available for 10 cups and 8 stems from 8 registries, totalling 552,148 cups and 727,447 stems. Follow-up was 1-20 years. Five-year CRRs pooled on all cups was 2.9% (95%CI 2.3 to 3.6) and on all stems 3.0% (95%CI 2.4 to 3.8). Homogenous (consistent) CRRs with respect to both, associated implant and country, were observed for 2 cups and 3 stems. Significant differences in CRR were identified in 1 cup by associated implant only, in 1 cup by registry only, and in 2 cups and 4 stems for both. Sparse data prevented evaluation of 4 cups and 1 stem.

**Conclusion:** Registries’ annual reports provide a large amount of publicly available information on CRRs of specific implants. These CRRs can be synthesized to improve the assessment of implant performance over time. Our CRR analysis represents a promising approach to detect implants with a consistent low- or high-risk pattern across registries.

## Introduction

Replacement of at least one component of a total hip arthroplasty (THA) - also called revision - is regarded as an important performance indicator of hip arthroplasty. Variation in revision risks may provide important information for clinicians to guide implant selection. Information on the risk of revision over time for a given cup, stem or cup/stem combination are potentially available from the peer-reviewed medical literature, acknowledged by the regulators as source of clinical evidence regarding medical devices (1), and from annual reports published by national/regional hip arthroplasty registries. The peer-reviewed literature may include early evidence on performance and safety of implants, that were part of obtaining the Conformité Européenne (CE) marking or shortly after market-entry. Registries on the other hand are monitoring the long-term, real-world performance of implants used in a country/region (2). The new European medical device regulation (MDR) reinforces the importance of post-market surveillance data and the role of registries (2). Registries often have procedures to benchmark revision rates at multiple time-points to rate safety and quality of individual implants or categories of implants (e.g. by type of fixation) (3, 4, 5). Because of security and data protection reasons, arthroplasty registries are reluctant or unable to share individual data to be pooled (6) and benchmarking and outlier detection is currently conducted at the national level. The lack of data sharing is a limitation for the detection of outliers and the assessment of an implants’ risk profile. Alternatively, meta-analyses based on aggregate data can be performed to combine registries data, but it has so far mostly been restricted to comparing categories of implants (7, 8, 9).

Combining revision rates of specific implant brands is highly desirable. It would allow testing the consistency of the revision results by examining them in different populations and settings, improve the precision of the estimated revision rate and increase the potential for stratified analyses. Finally, it would enable pooling of small numbers of implants from different registries and thus facilitate earlier detection of unsafe implants (3, 10). Analyses of combined revision rates for implants would be useful for many stakeholders including clinicians, hospitals, regulators, notified bodies, manufacturers, and health technology assessment agencies. Currently, publications in this area are limited. Hughes et al. published specific hip implant revision risks as reported by national and regional arthroplasty registries (11). However, the authors listed revision risks by implant and registry and did not present pooled results by implant.

The objective of this study is to systematically investigate the extent to which the cumulative risk of revision (CRR), for a random selection of currently used total hip stems and cups and for a frequently used cup and stem, is consistent across registries worldwide, or varies due to cup-stem combination (associated implant) and geographical location (registry country/region).

## Methods

The systematic review is reported according to the relevant items of the Preferred Reporting Items for Systematic Reviews and Meta-Analyses (PRISMA) statement (12) and it was registered on the Open Science framework (https://osf.io/6gmyx).

### Selection of implants

The selection of the assessed implants is described elsewhere (13, 14). Briefly, 10 cups and 10 stems were randomly selected from the Orthopaedic Data Evaluation Panel (ODEP) (15) and registry reports (combined list). Six cups were uncemented (Ana.Nova, Anexys, EcoFit, Exceed ABT, RM Pressfit Vitamys and Versafit Trio CC) and four cemented (Cenator, IP X-LINKed, Plasmacup SC and Polarcup). Nine stems were uncemented (Accolade II, Alloclassic, Avenir, BiContact, Collomis, Filler, MiniHip, Quadra H and Stelia stem) and one cemented (C-Stem AMT). Reported revision risks for these implants were searched in the registry reports. Two implants (cup Trident and stem Corail, both uncemented), that are frequently used internationally in current clinical practice were added to the search in registries to conduct further analyses like meta-regressions.

### Selection of registries and data collection

Registries were eligible if they provided in their annual report the cumulative risk of all-cause revision or the all-cause revision-free survival with 95% confidence intervals at any time point, for at least one of the 11 stems and 11 cups selected. Using the member list of the International Society of Arthroplasty Registries (ISAR) website (16) and a previous mapping of national and regional registries, the arthroplasty registries reporting annually the CRR and their 95% confidence interval (CI) by implant were identified (17). Registry country or region and the year of the latest annual report publication were collected. From registry reports published in English language, information was extracted for each cup-stem combination regarding the number of primary THAs recorded and the number of revisions for any cause, the corresponding CRRs and the 95%CI at all reported time-points. The initial data search was conducted in July 2021 and an update based on the latest reports has been made in November 2023.

### Statistical methods

CRRs were combined across registries by implant at follow-up of 1, 3, 5, 10 and 15 years using meta-analytic models with random effects and with the restricted maximum likelihood method. The random effect for the cups (respectively stems) associated with the assessed stem (respectively cup) was nested within the random effect for registry taking into account that the performance of specific cup-stem combinations might vary between registries. If the cup or the stem was reported by a single registry or with a single associated implant, the corresponding random effect was dropped from the model. For the meta-analyses, a complementary log-log transformation was applied to revision-free survivals, and standard errors were derived from the transformed 95%CI. Pooled estimates were back transformed and are presented as CRR. The presence of heterogeneity was investigated with Cochran’s Q test. For a given cup or stem, the relationship between the associated implants and the revision risk was investigated with a multiple meta-regression model with random effects and adjusted for country. With a complementary log-log link function, the exponential of the regression coefficient is an estimate of the hazard ratio (HR) (18). The residual heterogeneity was assessed (Cochran’s Q test and statistic I^2^). Statistical analyses were carried out with software R v4.0.2 (R Core Team (2022). R: A language and environment for statistical computing. R Foundation for Statistical Computing, Vienna, Austria. URL https://www.R-project.org/) and the package metafor v3.8-1 (19). All statistical tests were two-sided with a significance level of 0.05.

## Results

Overall, eight arthroplasty registries (six national and two regional) were included. Their list is presented in Appendix 1 and the flowchart of their selection in Appendix 2. Most of them are in Europe. Six additional registries reporting revision information by implant were identified but were not included because of a different definition of the outcome, a specific rather than the total population, incomplete reporting of CRR (only graphically reported or lack of confidence intervals) or reporting hazard ratios without the underlying rates.

Information in at least one of the eight registries was found for 10 of the 11 selected cups (90.9%) and eight of the stems (72.7%). Seven stems and five cups were assessed by more than one registry. For the randomly selected implants, the sample sizes were larger than 10,000 for six stems and three cups (Table 1). The largest sample sizes were for Versafit Trio CC cup (48,313 implants, four registries) and C-Stem AMT (70,823 implants, four registries). Length of follow-up varied considerably across implants (from 3 to 19 years for cups and from 7 to 15 years for stems) as well the number of associated implants (from 1 to 5 stems associated with a given cup and from 3 to 7 cups associated with a given stem). For the frequently used Trident cup and the Corail stem, that were added to the list of randomly selected implants, information was available for a much larger group of patients (391,475 and 427,313 prostheses), in six and eight registries, respectively, with a long follow-up and large numbers of associated implants.

**Table 1:** Implants reported by the registries.

|  | Registries |  |  |  |
| --- | --- | --- | --- | --- |
|  | Registries (n) | Primary THAs (n) | Associated implants (n) | Longest FU included (years) |
| Cup |  |  |  |  |
| Ana.Nova | 1 | 7,616 | 2 | 5 |
| AneXys | 1 | 3,072 | 2 | 3 |
| Cenator | 1 | 2,528 | 1 | 15 |
| EcoFit | 1 | 2,693 | 2 | 5 |
| Exceed ABT | 4 | 54,633 | 4 | 15 |
| IP X-LINKed | 2 | 2,099 | 1 | 5 |
| Plasmacup SC | 1 | 5,996 | 2 | 5 |
| Polarcup | 0 |  |  |  |
| RM Pressfit Vitamys | 3 | 33,723 | 4 | 10 |
| Versafit Trio CC | 4 | 48,313 | 6 | 10 |
| Trident* | 7 | 391,475 | 13 | 15 |
| Stem |  |  |  |  |
| Accolade II | 7 | 96,944 | 5 | 10 |
| Alloclassic | 3 | 36,835 | 7 | 15 |
| Avenir | 6 | 41,543 | 5 | 10 |
| BiContact | 1 | 16,454 | 4 | 5 |
| Collomis | 0 |  |  |  |
| Corail* | 8 | 427,313 | 14 | 15 |
| C-Stem AMT | 4 | 70,823 | 6 | 15 |
| Filler | 0 |  |  |  |
| MiniHip | 3 | 5,863 | 3 | 10 |
| Quadra H | 3 | 31,672 | 3 | 10 |
| Stelia stem | 0 |  |  |  |

\* selected as frequently used; THA=Total Hip Arthroplasty; FU=Follow-up

### Risk profile of the selected implants

The CRR pooled across all selected cups was 1.7% at one year of follow-up and increased to 2.5% at 3 years, 2.9% at 5 years, 4.0% at 10 years and 5.1% at 15 years (Figure 1A, Table 2). Various implant-specific patterns over time were observed: for some cups, 1-year CRR was low (e.g. Exceed ABT and RM Pressfit Vitamys) or the increase over time was low (e.g. IP X-LINKed and Plasmacup SC) (Figure 1A). Pooling the CRRs across registries confirmed the variability between implants (Table 2). At three years, the CRR was the highest (respectively lowest) for the EcoFit cup (respectively Cenator). For implants with long-term data available, differences became more apparent after 10 years reflecting that the increase of CRR over time was variable. Of the 3 cups with a long follow-up, the 15-year pooled CRRs varied between 2.6% (Exceed ABT) to 5.9% (Trident); the EcoFit cup with the highest 5-year revision risk had no longer time data available. The differences in CRRs between cups were detected by the meta-regression at 5 (p=0.025) and 10 years (p=0.009).

**Table 2:** Cumulative risks of revision (CRR) at 1, 3, 5, 10 and 15 years combined across registries by implant.

| Implant | Pooled CRR (95%CI) |  |  |  |  |
| --- | --- | --- | --- | --- | --- |
|  | 1 year | 3 years | 5 years | 10 years | 15 years |
| All cups | 1.69 (1.25 to 2.28) | 2.48 (1.92 to 3.21) | 2.93 (2.35 to 3.65) | 3.96 (2.86 to 5.49) | 5.09 (3.00 to 8.55) |
| Ana.Nova | 2.35 (1.26 to 4.37) | 2.94 (1.58 to 5.44) | 3.24 (1.61 to 6.50) |  |  |
| AneXys | 2.26 (0.89 to 5.67) | 2.66 (1.24 to 5.64) |  |  |  |
| Cenator | 0.64 (0.39 to 1.04) | 1.38 (0.99 to 1.93) | 2.05 (1.55 to 2.70) | 2.76 (2.15 to 3.53) | 4.30 (3.38 to 5.46) |
| EcoFit | 3.61 (2.10 to 6.17) | 4.41 (2.29 to 8.40) | 6.20 (3.94 to 9.69) |  |  |
| Exceed ABT | 1.18 (0.93 to 1.50) | 1.82 (1.42 to 2.32) | 2.04 (1.67 to 2.49) | 2.66 (1.97 to 3.57) | 2.63 (2.36 to 2.93) |
| IP X-LINKed | 1.88 (1.07 to 3.32) | 2.76 (1.94 to 3.93) | 3.58 (2.66 to 4.82) |  |  |
| Plasmacup | 2.02 (1.52 to 2.67) | 2.61 (2.22 to 3.08) | 2.76 (2.37 to 3.21) |  |  |
| Polarcup |  |  |  |  |  |
| RM Pressfit Vitamys | 1.83 (1.58 to 2.11) | 2.34 (1.96 to 2.78) | 2.78 (2.31 to 3.34) | 3.84 (2.81 to 5.25) |  |
| Trident | 1.67 (1.11 to 2.51) | 2.69 (1.90 to 3.80) | 3.29 (2.43 to 4.46) | 4.71 (3.07 to 7.19) | 5.94 (3.70 to 9.47) |
| Versafit CC Trio | 2.07 (1.57 to 2.72) | 2.73 (2.15 to 3.48) | 3.24 (2.56 to 4.10) | 6.15 (5.14 to 7.36) |  |
| All stems | 1.72 (1.19 to 2.50) | 2.49 (1.88 to 3.31) | 3.01 (2.40 to 3.77) | 4.49 (3.32 to 6.05) | 6.72 (3.23 to 13.72) |
| Accolade II | 1.91 (1.28 to 2.85) | 2.53 (1.84 to 3.47) | 2.83 (2.00 to 3.98) | 3.76 (1.20 to 11.49) |  |
| Alloclassic | 2.12 (1.20 to 3.72) | 3.05 (2.07 to 4.47) | 3.58 (2.61 to 4.89) | 4.34 (3.43 to 5.48) | 5.46 (3.45 to 8.59) |
| Avenir | 2.05 (1.21 to 3.47) | 2.36 (1.43 to 3.89) | 2.86 (1.91 to 4.26) | 3.65 (2.65 to 5.03) |  |
| BiContact | 3.04 (2.30 to 4.01) | 3.77 (2.83 to 5.02) | 4.35 (2.89 to 6.54) |  |  |
| C-Stem AMT | 1.05 (0.61 to 1.80) | 1.60 (1.02 to 2.51) | 1.88 (1.16 to 3.05) | 3.62 (1.66 to 7.78) | 6.32 (1.80 to 20.90) |
| Collomis |  |  |  |  |  |
| Corail | 1.50 (1.06 to 2.13) | 2.20 (1.66 to 2.90) | 2.72 (2.10 to 3.51) | 4.38 (3.04 to 6.30) | 7.70 (4.60 to 12.73) |
| Filler |  |  |  |  |  |
| MiniHip | 2.24 (1.35 to 3.72) | 3.06 (1.92 to 4.87) | 3.40 (2.18 to 5.30) | 3.17 (2.60 to 3.85) |  |
| Quadra H | 2.13 (1.75 to 2.60) | 3.09 (2.63 to 3.63) | 4.09 (3.09 to 5.40) | 6.12 (4.97 to 7.53) |  |
| Stelia stem |  |  |  |  |  |

**Figure 1:**
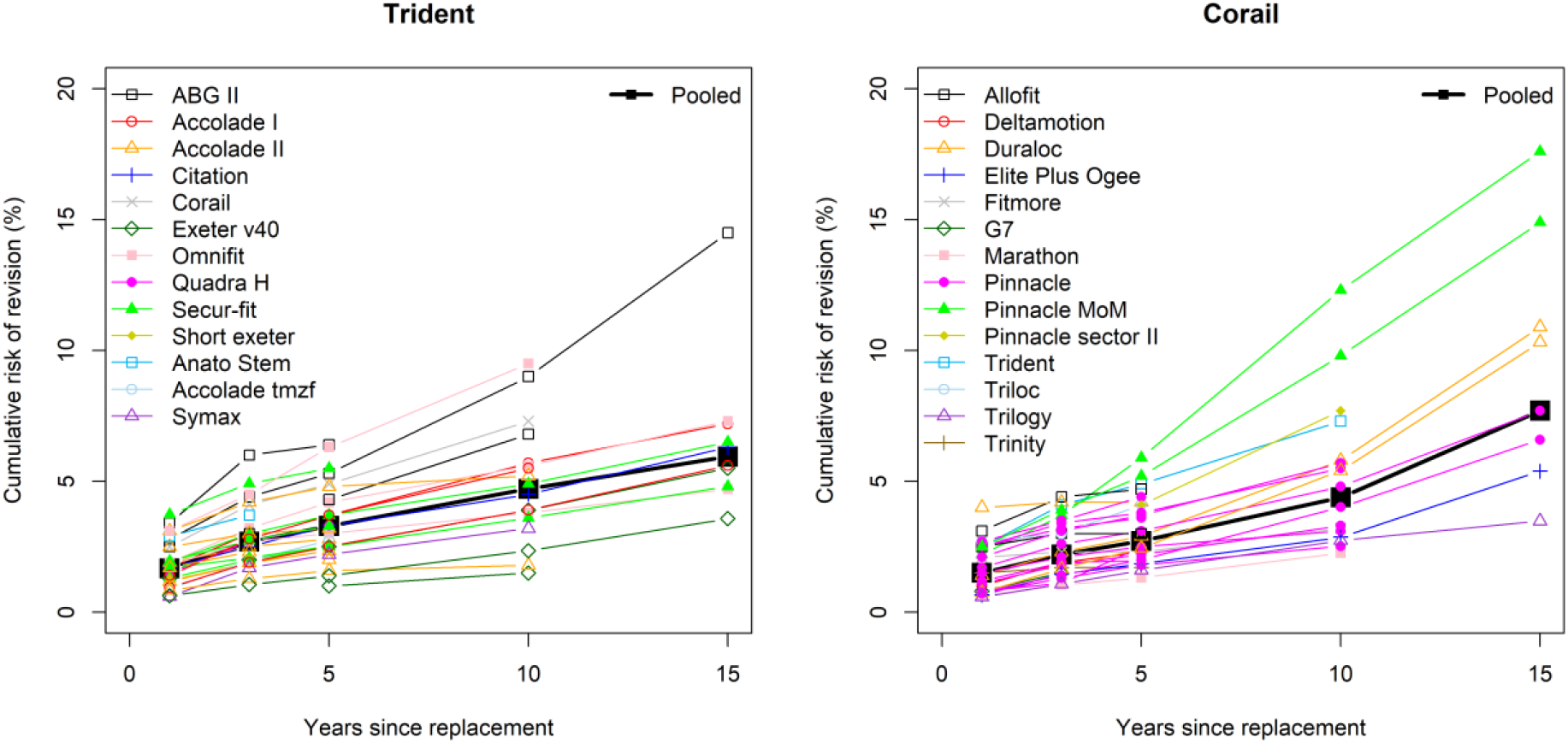
Overall pooled CRRs for cups (A; left panel) and for stems (B; right panel). The black squares represent the overall pooled CRR and the colored symbols represented the pooled CRR across registries for specific implants

For stems the CRRs pooled across all selected implants was 1.7% at one year of follow-up and increased to 2.5% at 3 years, 3.0% at 5 years, 4.5% at 10 years and 6.7% at 15 years (Figure 1B, Table 2). At 1 year, CRRs differed significantly (p=0.004) between implants at 1 year, e.g. three times higher for Bicontact than for C-stem (Figure 1B, Table 2). The increase over time also varied between implants. It either increased more strongly from the start, e.g. Quadra H, or after 5 to 10 years, e.g. C-stem and Corail. Of the three stems with long follow-up, 15-year pooled CRRs ranged from 5.5% (Alloclassic) to 7.7% (Corail). In addition to the 1-year difference, meta-regression detected further differences at 3 (p=0.046) and 15 years (p=0.011).

### Differences/Consistency by cup-stem combination and/or registries for the selected implants

Homogenous CRRs were observed for the cups IP X-LINKed and Plasmacup, and a single CRR over time was reported for Cenator. For three cups (Ana.Nova, AneXys and EcoFit) the source of the detected heterogeneity could not be investigated due to the small number of CRRs published. For the other cups, three differed significantly by associated implant (RM Pressfit Vitamys, Trident and Versafit CC Trio) and three by registry (Exceed ABT, Trident and Versafit CC Trio) (Appendix 3). For instance, CRRs for the RM Pressfit Vitamys were higher at 5 and 10 years when associated with Twinsys uncemented (pooled 10-year CRR 4.7%, 95%CI 4.1 to 5.4) than with Optimys (10-year CRR 2.9%, 95%CI 2.2 to 3.6) (Figure 2A), and the associated stems explained fully the apparent heterogeneity and the CRRs were homogeneous for each implant combination between the different registries. In contrast, for Versafit Trio CC the effect of the associated stems on the CRR was detected only at 5 years (Figure 2B, Appendix 3). At 10 years, although the results differed visually in particular for the Quadra C - only one registry reported it and with a large confidence interval - no difference was detected. Regarding the Trident, CRR was highest with the associated stems ABGII (pooled 10-year CRR 8.2%, 95%CI 6.4 to 10.6) and Omnifit (pooled 10-year CRR 6.5%, 95%CI 3.1 to 13.5) (Figure 2C) and lowest with Exeter v40 (pooled 10-year CRR 2.8%, 95%CI 1.7 to 4.4). The CRRs over time by associated stem are displayed in Appendix 4 for the other cups.

**Figure 2:**
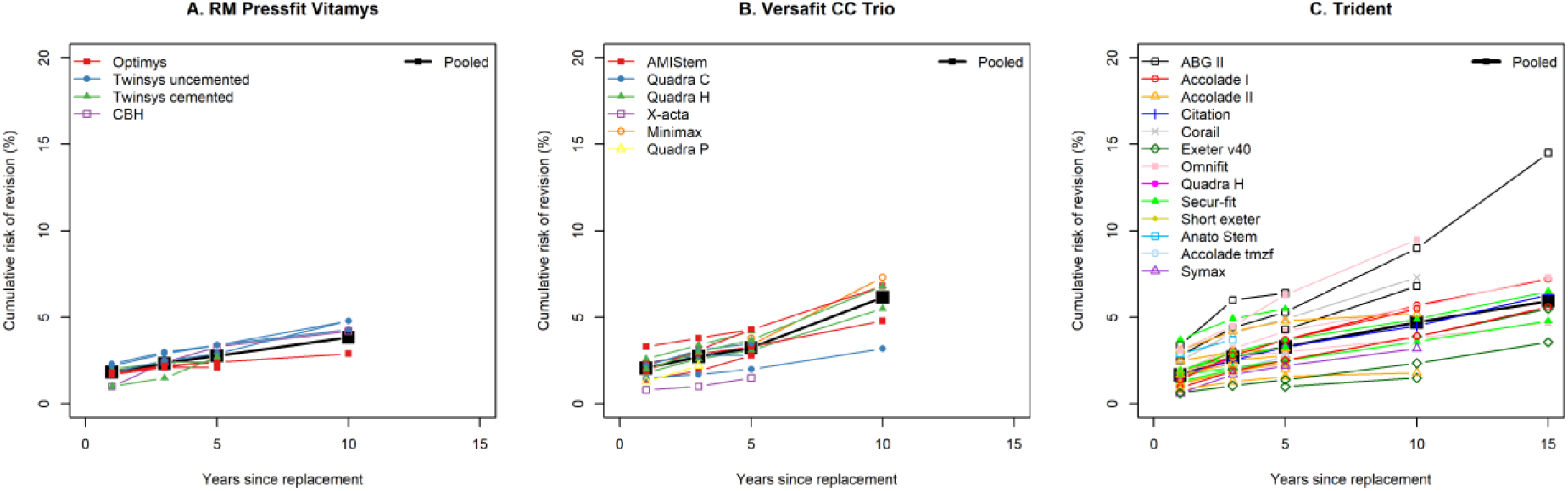
Pooled CRRs (black squares) and CRRs published in annual registries reports for the cups A) RM Pressfit Vitamys, B) Versafit CC Trio and C) Trident depending on stem combination (coloured symbols)

The meta-regression model for Trident for the 5-year CRR confirmed that the CRR adjusted for the registry country was higher for the associated stems ABGII (HR=2.4), Corail (HR=2.2) and Omnifit (HR=1.8) than with Exeter v40, and that the CRR adjusted for the associated stem was higher in Finland (HR=2.9), the Netherlands (HR=2.0) and Germany (HR=1.9) than in the United Kingdom (UK) (Table 3). The residual heterogeneity, that is the heterogeneity unexplained by registry or associated implant, remained high for the Trident cup (Appendix 3).

**Table 3:** Multivariable model for the 5-year cumulative risk of revision with the cup Trident.

|  | N implants | Adjusted HR<br>(95%CI) | P-value |
| --- | --- | --- | --- |
| Registry country/region |  |  |  |
| England, Wales | 196,048 | 1 (ref.)* | **0.006\*\*** |
| Australia | 141,187 | 1.51 (1.06 to 2.15) | 0.021 |
| Finland | 1,877 | 2.95 (1.59 to 5.47) | 0.001 |
| Germany | 6,034 | 1.90 (1.12 to 3.21) | 0.017 |
| Emilia Romagna, Italy | 1,475 | 1.13 (0.59 to 2.17) | 0.711 |
| Michigan | 27,856 | 1.59 (1.01 to 2.51) | 0.045 |
| Netherlands | 11,179 | 1.98 (1.21 to 3.23) | 0.006 |
| Associated stem |  |  |  |
| Exeter v40 | 233,284 | 1 (ref.)* | **0.005\*\*** |
| ABG II | 3,460 | 2.43 (1.45 to 4.07) | 0.001 |
| Accolade I | 44,196 | 1.54 (0.99 to 2.38) | 0.054 |
| Accolade II | 66,441 | 1.11 (0.73 to 1.70) | 0.614 |
| Accolade tmzf | 913 | 1.17 (0.54 to 2.52) | 0.696 |
| Citation | 1,147 | 1.48 (0.76 to 2.88) | 0.243 |
| Corail | 588 | 2.22 (1.07 to 4.59) | 0.031 |
| Omnifit | 5,768 | 1.77 (1.10 to 2.85) | 0.019 |
| Quadra H | 712 | 1.48 (0.73 to 3.03) | 0.28 |
| Secur-fit | 22,987 | 1.46 (0.93 to 2.30) | 0.102 |
| Short Exeter | 4,087 | 1.12 (0.59 to 2.11) | 0.728 |
| Symax | 2,073 | 0.75 (0.35 to 1.62) | 0.464 |

\*Registry country/region and associated stem with the largest sample size selected as category of reference
\*\*Overall p-values

Homogenous CRRs with respect to associated implant and country were observed for the stem Quadra H (Figure 3A), Minihip, and Avenir. For one stem (Bicontact) the source of the detected heterogeneity could not be investigated due to the small number of CRRs published. All other stems differed significantly by associated cup. For four stems (Accolade II, Alloclassic, C-stem AMT, Corail) the results differed significantly by registry (Appendix 3). The patterns of CRRs for specific stems over time could vary depending on the associated cup. For instance, the CRRs were higher for the C-Stem AMT when associated with the Duraloc (15-year CRR 12.3% 95%CI 10.0 to 15.1) The associated cups Marathon, Charnley Elite plus and Elite plus ogee presented similar patterns with lower CRRs (Figure 3B). For the frequently used Corail stem, the CRR at long follow-up reached a high level when associated with Duraloc (pooled 15-year CRR 10.4%, 95%CI 9.5 to 11.5) and with Pinnacle MoM (pooled 15-year CRR 16.6%, 95%CI 14.3 to 19.4) (Figure 3C). Similar trajectories of these two implants up to 15 years were found in two different registries. The CRR differences between the associated cups became more visible after 10 years. CRRs over time by associated cup and by registry location are displayed in Appendix 5 for the other stems.

**Figure 3:**
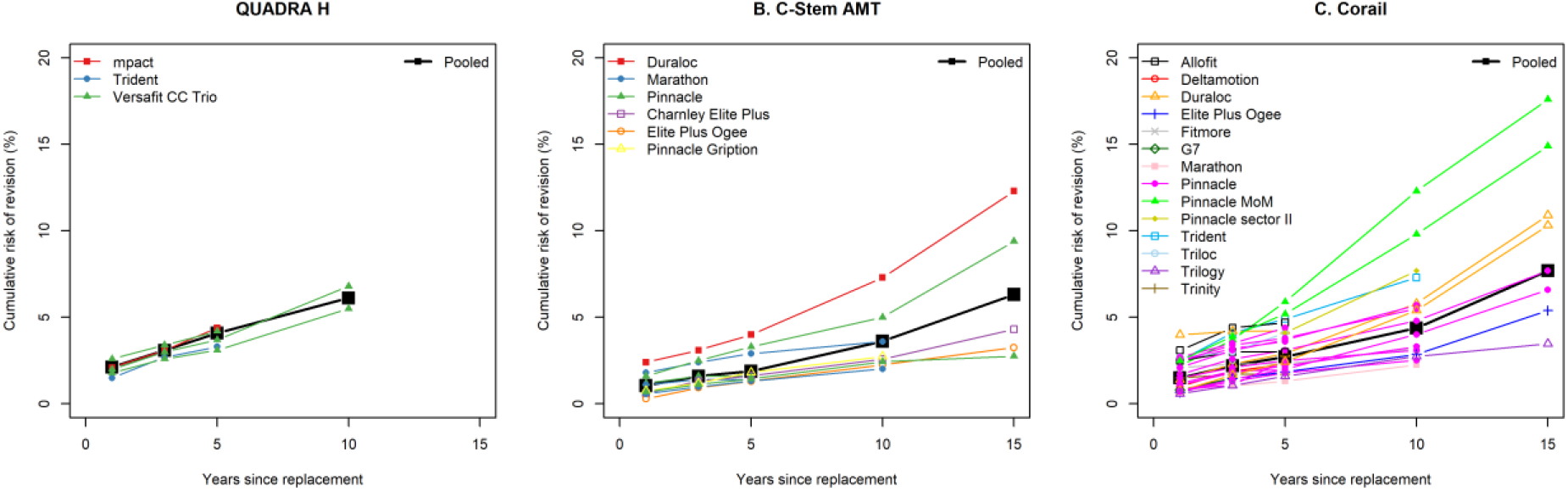
Pooled CRRs (black squares) and CRRs published in annual registries reports for the stems A) QUADRA H, B) C-Stem AMT and C) Corail depending on cup combination (coloured symbols)

The patterns of CRRs for specific stems over time could vary depending on the associated cup. For instance, the CRRs were higher for the C-Stem AMT when associated with the Duraloc (15-year CRR 12.3% 95%CI 10.0 to 15.1) The associated cups Marathon, Charnley Elite plus and Elite plus ogee presented similar patterns with lower CRRs (Figure 3B). For the frequently used Corail stem, the CRR at long follow-up reached a high level when associated with Duraloc (pooled 15-year CRR 10.4%, 95%CI 9.5 to 11.5) and with Pinnacle MoM (pooled 15-year CRR 16.6%, 95%CI 14.3 to 19.4) (Figure 3C). Similar trajectories of these two implants up to 15 years were found in two different registries. The CRR differences between the associated cups became more visible after 10 years. CRRs over time by associated cup and by registry location are displayed in Appendix 5 for the other stems.

The meta-regression model on the 5-year CRR adjusted for the registry country/region showed that the CRR was lower with the Marathon (HR=0.7) and Trinity (HR=0.5) than with the Pinnacle (Table 4). It also showed that the CRR was higher in Australia (HR=1.5), Finland (HR=1.7) Germany (HR=2.0) and Switzerland (HR=1.6) than in the UK. The residual heterogeneity, i.e. the heterogeneity unexplained by registry country or associated implant, remained high for the Corail stem (Appendix 3).

**Table 4:** Multivariable model for the 5-year cumulative risk of revision with the stem Corail.

|  | 5-year CRR |  |  |
| --- | --- | --- | --- |
|  | N implants | Adjusted HR<br>(95%CI) | P-value |
| Registry country/region |  |  |  |
| England, Wales | 241,750 | 1 (ref.)* | **<0.001\*\*** |
| Australia | 68,443 | 1.55 (1.16 to 2.08) | 0.003 |
| Finland | 2,184 | 1.68 (1.12 to 2.53) | 0.012 |
| Germany | 42,565 | 1.98 (1.50 to 2.62) | <0.001 |
| Emilia Romagna, Italy | 1,065 | 1.83 (0.61 to 5.51) | 0.283 |
| Michigan | 3,148 | 0.99 (0.64 to 1.54) | 0.975 |
| Netherlands | 43,606 | 1.16 (0.85 to 1.58) | 0.357 |
| Switzerland | 23,931 | 1.56 (1.15 to 2.10) | 0.004 |
| Associated cup |  |  |  |
| Pinnacle | 385,081 | 1 (ref.)* | **0.005\*\*** |
| Allofit | 1,987 | 0.91 (0.61 to 1.35) | 0.635 |
| Deltamotion | 1,353 | 0.75 (0.44 to 1.28) | 0.291 |
| Duraloc | 6,008 | 1.10 (0.83 to 1.45) | 0.499 |
| elite plus ogee | 3,188 | 0.93 (0.60 to 1.44) | 0.752 |
| Fitmore | 514 | 0.75 (0.37 to 1.50) | 0.414 |
| Marathon | 19,719 | 0.66 (0.45 to 0.96) | 0.029 |
| Pinnacle MoM | 2,148 | 1.76 (1.22 to 2.55) | 0.002 |
| Pinnacle Sector II | 719 | 1.14 (0.35 to 3.75) | 0.827 |
| Trident | 588 | 1.62 (0.91 to 2.88) | 0.102 |
| Triloc | 842 | 1.05 (0.60 to 1.85) | 0.854 |
| Trilogy | 3,319 | 0.80 (0.52 to 1.26) | 0.339 |
| Trinity | 1,226 | 0.55 (0.31 to 0.99) | 0.048 |

\*Registry country/region and associated cup with the largest sample size selected as category of reference
\*\*Overall p-values

## Discussion

This review shows that graphical representations and quantitative syntheses of reported CRRs, by applying statistical methods for meta-analyses, can be used to facilitate the assessment of implant risk profile over time from multiple registries. We identified differences in performance of selected implants and more importantly in the performance of cup-stem combinations. The other source of heterogeneity was the registry, which can be stratified by and adjusted for in meta-regression in case of sufficient data. Although the amount of data published by registries is an advantage, it makes appraisal and synthesis of the risk of revision of the many implants challenging; the graphical representation and the meta-analyses of the CRRs over time we propose here are helpful to identify global patterns.

The CRRs varied importantly by implant. For instance, for some cups, the 10-year CRRs were around 3% and around 7% for others. The difference in patterns mainly became readable after 5 years.

The overall pooled CRRs at 5 and 10 years in our study (2.9% and 4.0% for cups; 3.0% and 4.5% for stems) were comparable to the all-construct survivorship estimates reported by Paxton et al. in 2019 (Sweden 5-year 97.8% and 10-year survival 95.8%, United States 5-year 97.0% and 10-year 95.2%, and Australia 5-year 96.3% and 10-year 93.5%) (20). The pooled all-construct survivorship at 15 years derived from registry data from Australia, Denmark, Finland, New Zealand, Norway, and Sweden including patients operated up to 2017 was 89.4% (95% CI 89.2–89.6) (8). Our pooled CRRs of 5.1% for cups and 6.7% for stems at 15 years including THAs operated up to 2022 were lower, which might be related to the inclusion of more recent implants and the fact that differences become more apparent at longer follow-up. An overall decrease in revision rates by year since 2008 has been reported by the National Joint Registry (21) and the Dutch Arthroplasty Register (LROI) (22).

### Limitations

For some of the implants (4 cups and 1 stem) it was not possible to investigate the potential sources of heterogeneity due to the low number of registries reporting CRRs. The inclusion of additional registries would increase the amount of data and allow going further in their assessment. The potential sources of heterogeneity we investigated were limited to the associated implants and the country but the heterogeneity in CRRs may also be due to inaccurate or insufficiently granular implant labelling leading to grouping of heterogenous implants under one name, differences in case mix between countries and implant combinations, differences in associated bearing surface, differences in surgical techniques used (especially surgical approach) and health system/policy factors that drive the decision to revise among others (20).

The meta-analytic approach we propose is promising but methodological improvements are necessary. Conducting meta-analyses independently at each time point multiplies statistical testing and may lead to inconsistent pooled CRRs (e.g. decreasing pooled CRRs over time) because the documented time points vary across registries. To address these issues, a statistical method is needed to assess pooled CRRs over time with a single model.

This review has several limitations related to the reporting of CRRs and to the implant identification/labelling in the annual reports of registries. Since CRRs were not available by age and other risk factors for revision, it was not possible to conduct meta-analyses stratified on those factors and to draw fine tuned conclusions on the risk profile of implants accounting for the risk profile of patients. In addition, case mix of patients between registries, between implants within registries and between implant combinations is a potential source of confounding. The random effects introduced in the statistical models may imperfectly account for the case mix. The differences in CRRs between implants or implant combinations cannot be interpreted as causal relationships, although consistent patterns in several registries suggest causality. In addition, the role of the bearing surface could not be investigated here since CRRs of implant combinations were not systematically reported by bearing surface. Potential implant misclassification caused by an imprecise or insufficient implant identification/labelling in the annual reports is also possible (e.g. Corail details regarding collared or non-collared stem not always reported; different versions of associated cup Pinnacle not systematically detailed). Such misclassifications may produce an excess of heterogeneity, which would be addressed with standardized and more granular implant labelling.

### Strengths

Our study highlights the enormous value of prospective nation- or region-wide data collection with high a coverage (94%-99% for the established registries) and representativity of the sample, with harmonized baseline and outcomes data collection (23) and transparent public reporting of implant performance. Half of the registries reported the outcome of interest as CRRs with 95%CIs, which made it possible to pool the data with meta-analytic methods. The graphical representation of CRRs over time by implant combination and the meta-analyses allows for appraisal of risk patterns and testing of their consistency across different registries. This is helpful to identify not only high-risk implants but also implants or implant combinations showing a consistent low-risk pattern that can be used as standard comparators.

### Perspectives

Despite the limitations and the need for additional methodological developments, the approach we propose was able to identify different patterns in cup-stem combination CRRs in particular from 5 years on. It is promising for the early detection of outliers. This would be even more efficient if the number of registries reporting CRRs with confidence intervals by implant combination increases. For this, further efforts in harmonized registry reporting are needed. Our approach to synthesising survival outcomes is not limited to orthopedic implants but can also be applied to assess the risk profile of implants across countries in other medical areas (e.g. cardiology).

Finally, CRR variability between registries as shown in this study calls for rethinking the process of international benchmarking. The observed CRRs depend not only on the intrinsic performance of the implants but also on the population, surgery-related factors and on country-/region-specific health care practices and access to care. Thus, rating implants in all registries using the same absolute values/limits seems suboptimal. Within-registry benchmarking assessing the specific implant’s performance against a comparator group (e.g. all contemporary implants in the registry) and examining in a second step whether there is consistency between registries in the implant’s risk pattern over time and in its comparative performance might be a way forward worth investigating.

## Conclusion

Registries provide a large amount of publicly available information on specific implant CRRs that can be graphically represented and synthesized to investigate the risk profile of implants depending on the associated implant and country. The approach we proposed is promising to detect implant combinations with a consistently low- or high-risk pattern across registries.

## Data Availability

All data analyzed in this studies are publicly available in the annual reports of arthroplasty registries.

