## Appendix 1 for "Cumulative risk of revision after primary total hip arthroplasty in registries: Systematic review and meta-analysis of selected hip stems and cups"

Appendix 1: Total hip arthroplasty registries included in the review

| Name of registry | Type | Established | Link to annual reports available prior to December 8. 2023 |
|---|---|---|---|
| Australian Orthopaedic Association National Joint Replacement Registry (AOANJRR) | National | 1999 | https://aoanjrr.sahmri.com/annual-reports-2023 |
| Dutch Arthroplasty Register (LROI) | National | 2007 | https://www.lroi-report.nl/ Report 2023 |
| Endoprothesenregister Deutschland (EPRD) | National | 2012 | https://www.eprd.de/en/downloads/reports (EPRD Annual Report 2022) |
| Finnish Arthroplasty Register (FAR) | National | 1980 | https://www2.thl.fi/endo/report/#index (Dynamic site visited on November 2023) |
| Michigan Arthroplasty Registry Collaborative Quality Initiative (MARCQI) | Regional | 2011 | https://marcqi.org/dev/wp-content/uploads/2023/01/2022-REPORT-1-23-2023.pdf |
| National Joint Registry (NJR), United Kingdom | National | 2002 | https://reports.njrcentre.org.uk/Portals/0/PDFdownloads/NJR%2020th%20Annual%20Report%202023.pdf |
| Register of Orthopaedic Prosthetic Implants – RIPO Emilia Romagna | Regional | 2000 | https://ripo.cineca.it/authzssl/Reports.html (Annual report 2018 Regione Emilia – Romagna in English) |
| Swiss arthroplasty registry (SIRIS) | National | 2012 | https://www.siris-implant.ch/fr/Downloads&category=16 SIRIS Report 2023 |
