## Supplementary figures and images for "Cumulative risk of revision after primary total hip arthroplasty in registries: Systematic review and meta-analysis of selected hip stems and cups"

### Appendix 2

Appendix 2: Flowchart for the selection of total hip arthroplasty registries

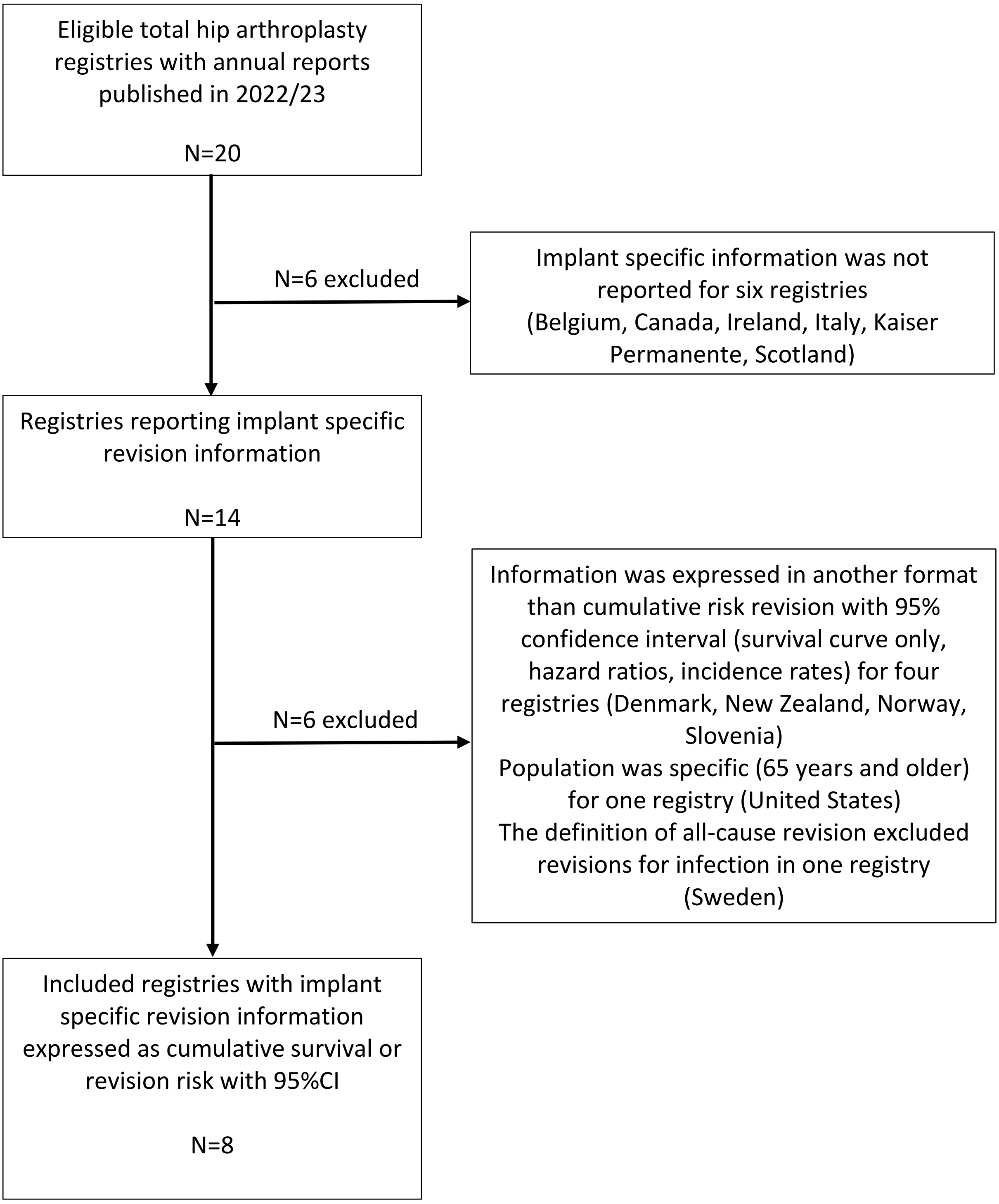
