## Appendix 3 for "Cumulative risk of revision after primary total hip arthroplasty in registries: Systematic review and meta-analysis of selected hip stems and cups"

Appendix 3: Heterogeneity, statistical significance of the association between country/associated implant and CRR, and the residual heterogeneity*

| | P-value for heterogeneity | | | | | P-value for associated implant** | | | | | P-value for country registries** | | | | | $I^2$ (%) for residual heterogeneity*** | | | | |
|---|---|---|---|---|---|---|---|---|---|---|---|---|---|---|---|---|---|---|---|---|
| **Implant** | 1y | 3y | 5y | 10y | 15y | 1y | 3y | 5y | 10y | 15y | 1y | 3y | 5y | 10y | 15y | 1y | 3y | 5y | 10y | 15y |
| Cups | | | | | | | | | | | | | | | | | | | | |
| Ana.Nova | 0.006 | 0.005 | 0.001 | | | | | | | | | | | | | | | | | |
| AneXys | <0.001 | 0.001 | | | | | | | | | | | | | | | | | | |
| EcoFit | 0.051 | 0.006 | | | | | | | | | | | | | | | | | | |
| Exceed ABT | 0.006 | <0.001 | <0.001 | <0.001 | | 0.512 | 0.958 | 0.893 | 0.899 | | 0.161 | 0.012 | 0.102 | 0.031 | | | 49.3 | | 77.3 | |
| IP X-LINKed | 0.084 | 0.219 | 0.268 | | | | | | | | | | | | | | | | | |
| Plasmacup | 0.4370 | 0.756 | 0.836 | | | | | | | | | | | | | | | | | |
| RM Pressfit Vitamys | 0.185 | 0.109 | 0.033 | 0.010 | | 0.070 | 0.014 | 0.002 | 0.002 | | 0.367 | 0.454 | 0.393 | 0.768 | | | 0.0 | 0.0 | 0.0 | |
| Trident | <0.001 | <0.001 | <0.001 | <0.001 | <0.001 | 0.075 | 0.082 | 0.005 | 0.007 | 0.001 | 0.004 | 0.005 | 0.006 | 0.012 | 0.089 | 90.1 | 90.7 | 90 | 95.6 | 93.4 |
| Versafit CC Trio | <0.001 | <0.001 | <0.001 | 0.021 | | 0.175 | 0.117 | 0.033 | 0.344 | | <0.001 | <0.001 | 0.005 | 0.333 | | 29.2 | 37.6 | 30.7 | | |
| Stems | | | | | | | | | | | | | | | | | | | | |
| Accolade II | <0.001 | <0.001 | <0.001 | <0.001 | | 0.007 | 0.095 | 0.004 | <0.001 | | <0.001 | <0.001 | <0.001 | <0.001 | | 0.0 | 0.0 | 0.0 | 0.0 | |
| Alloclassic | <0.001 | <0.001 | <0.001 | <0.001 | 0.0001 | 0.556 | 0.001 | 0.170 | 0.065 | | 0.014 | <0.001 | 0.040 | 0.690 | | 81.3 | 24.8 | 69.7 | | |
| Avenir | <0.001 | <0.001 | <0.001 | 0.018 | | 0.315 | 0.414 | 0.068 | 0.514 | | 0.046 | 0.355 | 0.051 | 0.282 | | 88.2 | | | | |
| BiContact | <0.001 | <0.001 | <0.001 | | | | | | | | | | | | | | | | | |
| C-Stem AMT | <0.001 | <0.001 | <0.001 | <0.001 | <0.001 | 0.010 | 0.262 | 0.304 | 0.031 | 0.7049 | <0.001 | <0.001 | <0.001 | <0.001 | <0.001 | 0.0 | 0.0 | 0.0 | 0.0 | 56.5 |
| Corail | <0.001 | <0.001 | <0.001 | <0.001 | <0.001 | 0.552 | 0.216 | 0.005 | 0.083 | <0.0001 | <0.001 | <0.001 | <0.001 | 0.7565 | 0.001 | 57.3 | 75.2 | 75.0 | 93.1 | 0.0 |
| MiniHip | 0.002 | 0.002 | 0.003 | 0.316 | | 0.532 | 0.266 | 0.186 | | | 0.109 | 0.423 | 0.306 | | | | | | | |
| Quadra H | 0.008 | 0.047 | 0.002 | 0.069 | | 0.630 | 0.896 | 0.467 | | | 0.022 | 0.326 | 0.812 | | | 18.9 | | | | |

*: heterogeneity that was not explained by the registry nor the associated implant

**: the associations between associated implant (respectively registry) and CRR were assessed only for implants reported with at least three combinations (respectively three registries)

***: the residual heterogeneity was assessed only if an association with the associated implants or with the registries was detected
