## Appendix 4 for "Cumulative risk of revision after primary total hip arthroplasty in registries: Systematic review and meta-analysis of selected hip stems and cups"

Appendix 4: CRRs for cups by registry and associated stem. Black squares represent the pooled CRRs independently of the associated stem and coloured symbols represent the associated stems.

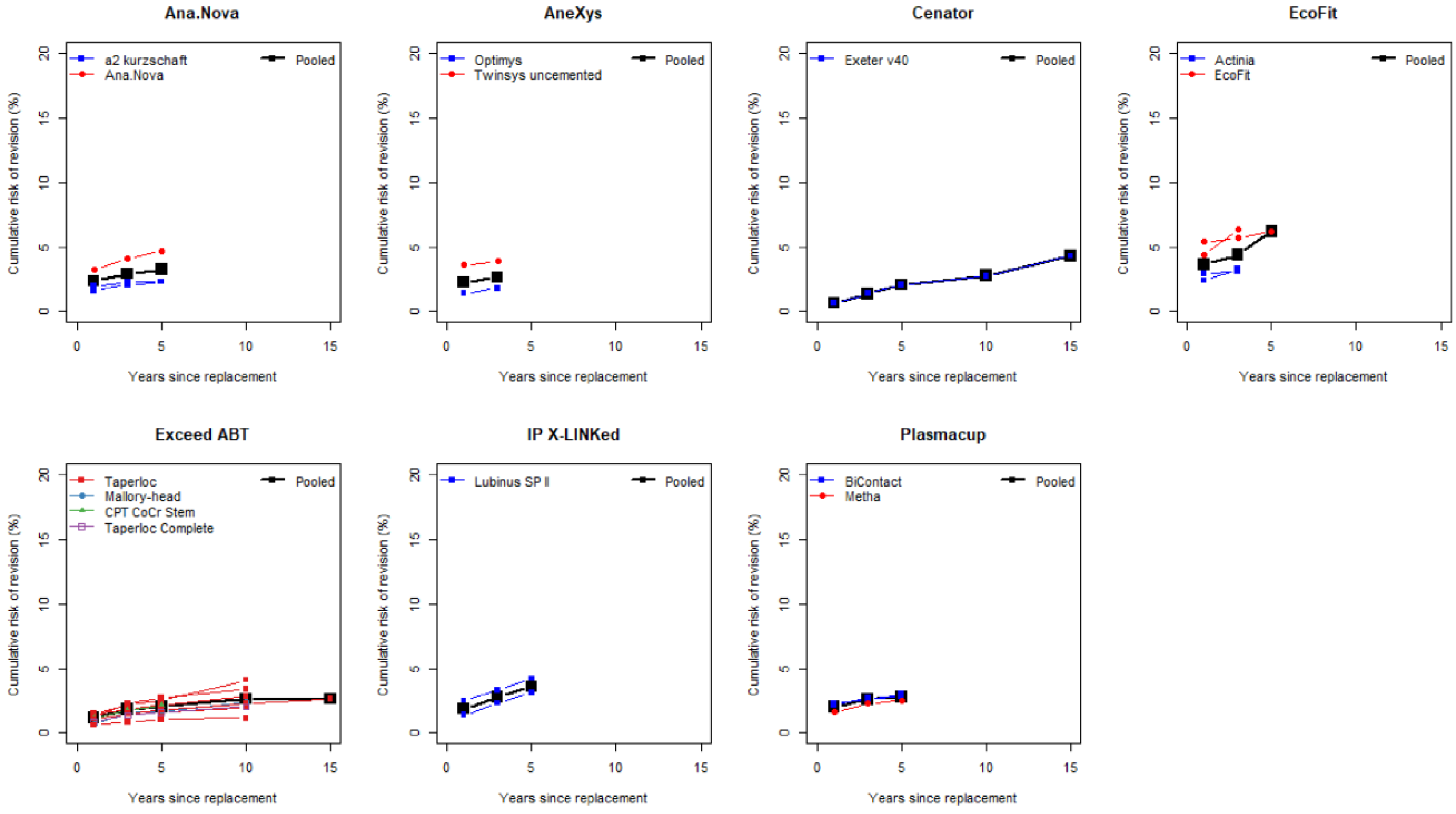
