## Appendix 5 for "Cumulative risk of revision after primary total hip arthroplasty in registries: Systematic review and meta-analysis of selected hip stems and cups"

Appendix 5: CRRs for stems by registry and associated cup. Black squares represent the pooled CRRs independently of the associated cups and coloured symbols represent the associated cups.

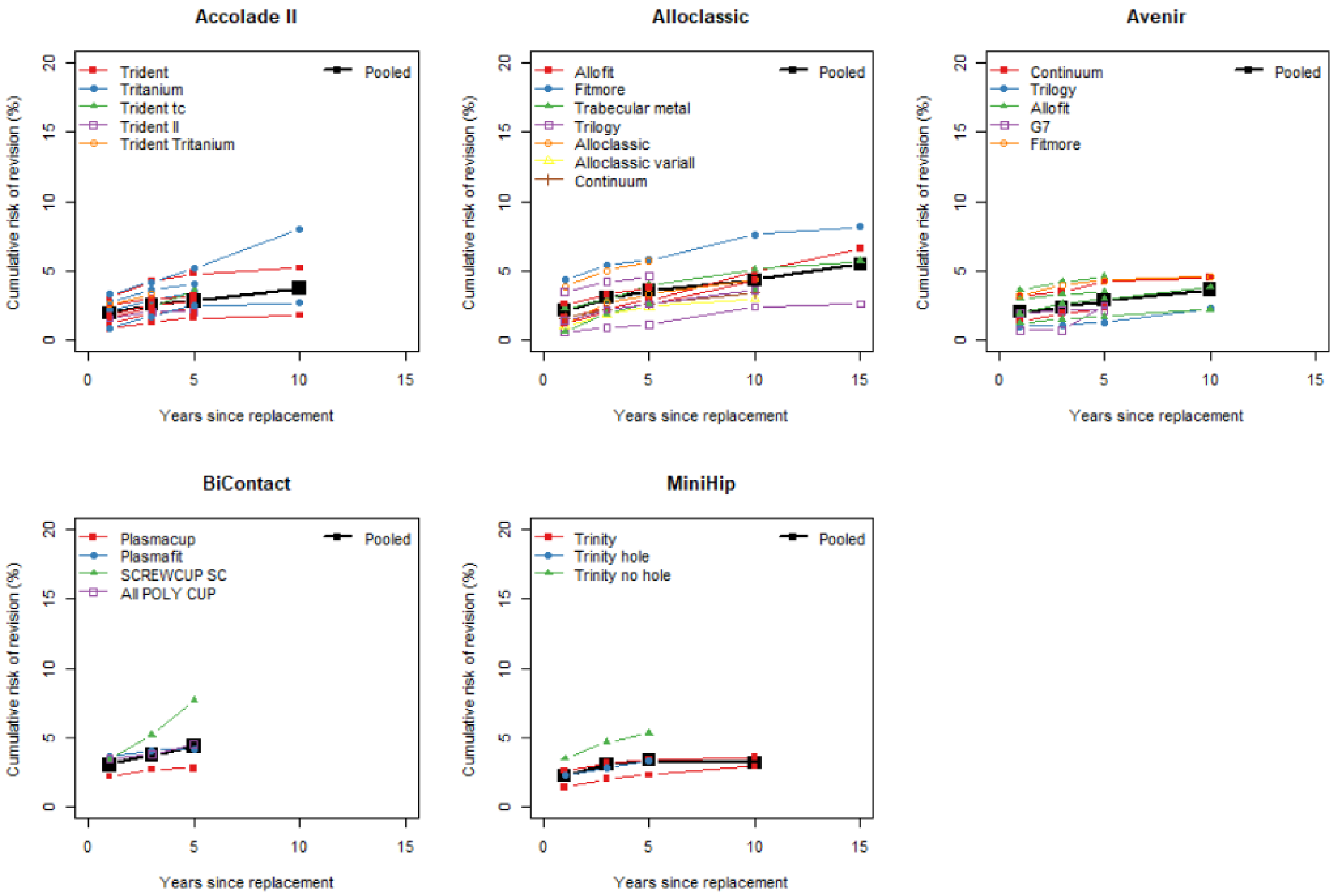
